# Sputnik-V reactogenicity and immunogenicity in the blood and mucosa: a prospective cohort study

**DOI:** 10.1101/2022.01.26.22269729

**Authors:** Sergey Yegorov, Irina Kadyrova, Baurzhan Negmetzhanov, Yevgeniya Kolesnikova, Svetlana Kolesnichenko, Ilya Korshukov, Yeldar Baiken, Bakhyt Matkarimov, Matthew S. Miller, Gonzalo H. Hortelano, Dmitriy Babenko

## Abstract

**Background:** Sputnik-V (Gam-COVID-Vac) is a heterologous, recombinant adenoviral (rAdv) vector-based, COVID-19 vaccine now used in >70 countries. Yet there is a shortage of data on this vaccine’s performance in diverse populations. Here, we performed a prospective cohort study to assess the reactogenicity and immunologic outcomes of Sputnik-V vaccination in a multiethnic cohort from Kazakhstan.

**Methods:** COVID-19-free participants (n=82 at baseline) were followed at day 21 after Sputnik-V dose 1 (rAd5) and dose 2 (rAd26). Self-reported local and systemic adverse events were captured using questionnaires. Blood and nasopharyngeal swabs were collected to perform SARS-CoV-2 diagnostic and immunologic assays.

**Findings:** Of the 73 and 70 participants retained post-dose 1 and 2, respectively, most (>50%) reported mild-to-moderate injection site or systemic reactions to vaccination; no severe or potentially life-threatening conditions were reported. dose 1 appeared to be more reactogenic than dose 2, with fatigue and headache more frequent in participants with prior COVID-19 exposure. After dose 2 nausea was more common in subjects without prior COVID-19. The combined S-IgG and S-IgA seroconversion rate was 97% post-dose 1, remaining the same post-dose 2. The proportion of participants with detectable virus neutralization titers was 83% post-dose 1’, and increased to 98% post-dose 2’, with the largest relative increase observed in participants without prior COVID-19 exposure. Nasal S-IgG and S-IgA increased post-dose 1, while the boosting effect of dose 2 on mucosal S-IgG, but not S-IgA, was only observed in subjects without prior COVID-19. Systemically, vaccination reduced serum levels of growth regulated oncogene (GRO), which correlated with an elevation in blood platelet count.

**Interpretation:** Sputnik-V dose 1 elicited both blood and mucosal SARS-CoV-2 immunity, while the immune boosting effect of dose 2 was minimal, suggesting that adjustments to the current vaccine dosing regimen may be necessary to optimize immunization efficacy and cost-effectiveness. Although Sputnik-V appears to have a reactogenicity profile similar to that of other COVID-19 vaccines, the observed alterations to the GRO/platelet axis call for further investigation of Sputnik V effects on systemic immunology.

**Funding:** Ministry of Education and Science of the Republic of Kazakhstan.

## INTRODUCTION

Sputnik-V, also known as Gam-COVID-Vac, is a recombinant adenovirus vector-based vaccine against COVID-19, developed by in Russia ^1,2^. The vaccine has a heterologous prime-boost regimen and is typically administered in two doses containing rAd26 as a prime and rAd5 as a boost, with a 21-day interval between the doses. The initial clinical trials demonstrated that Sputnik-V vaccination resulted in complete prevention of severe COVID-19 and was 91.6% effective against symptomatic SARS-CoV-2 infection ^1,2^.

Sputnik-V has been approved for use in over 70 countries ^3^; despite such broad geographic distribution, there is still a dearth of data describing the performance of the vaccine. This lack of rigorous data has precluded major regulatory agencies and the World Health Organization from granting Sputnik-V emergency use approval ^4^. The studies conducted after the original Phase I-III trials have indicated that, like the COVID-19 vaccines approved in the West, Sputnik-V is safe and elicits robust titers of binding and neutralizing antibodies (Ab) against SARS-CoV-2 ^5–18^. However, more detailed data on the reactogenicity and immunogenicity of Sputnik-V are urgently needed to not only facilitate vaccine deployment, but also to guide population-specific guidelines regarding matters such as the timing of prime-boost doses, which remains a focus of debate ^16–18^. Importantly, no data are currently available on the antibody isotypes, other than IgG, elicited by Sputnik-V in the blood or mucosa. A recent study of mRNA vaccination, for example, indicates that breakthrough COVID-19 is associated with lower serologic titers of S/RBD-specific IgA, but not IgG ^19^, thus emphasizing the need to include IgA in vaccine immunogenicity assessments.

The first confirmed cases of COVID-19 in Kazakhstan were identified in mid-March 2020 ^20^. Subsequent rapid community spread led to substantial morbidity and mortality in the country ^20–22^. Sputnik-V vaccination roll-out in Kazakhstan began in February 2021, with several other COVID-19 vaccines later added to the national vaccination portfolio. This early 2021 Sputnik-V rollout among public employees in Karaganda, Kazakhstan presented us with an opportunity to conduct a prospective study of self-reported adverse reactions and immunologic responses after Sputnik-V vaccination in a cohort with and without prior history of COVID-19 exposure.

## METHODS

### Study population and design

This registered clinical trial (ClinicalTrials.gov #NCT04871841) was conducted in Karaganda, the capital of the Karaganda region located in Central Kazakhstan, where serologically confirmed SARS-CoV-2 exposure exceeds 63% ^23^. The initial participant screening occurred in April-May 2021 at the Karaganda Medical University COVID-19 vaccination site. Consenting, asymptomatic adults, who had not previously received a COVID-19 vaccine, were invited to participate. Exclusion criteria were presence of respiratory symptoms or laboratory-confirmed COVID-19 diagnosis within two weeks prior to the study. Short questionnaires addressing the participants’ demographic background and recent history of COVID-19 exposure were administered. At follow-up, participants were screened for respiratory symptoms and tested for COVID-19 breakthrough infections using SARS-CoV-2 PCR and nucleocapsid protein (NCP) ELISA. Participants, who were not eligible due to the presence of respiratory conditions, were referred to the clinic physician for counselling and treatment. We used earlier studies ^2,16,18^ as reference point when establishing the study sample size, which was ultimately constrained by the study budget.

### Vaccination procedures

The vaccine was administered after collection of biological samples according to the National Ministry of Healthcare guidelines by the clinic staff. Specifically, a 0.5 ml dose of Sputnik-V (sourced from Moscow, Russia) containing 1± 0.5 ×10^11^ rAd particles was injected into the deltoid muscle. Doses 1 and 2 consisted of rAd26 and rAd5, respectively.

### Adverse event data collection and reporting

Standardized questionnaires regarding the solicited adverse events (AE), pre-defined as local (at the injection site) or systemic were administered at the 21-day post-vaccination follow-up visits. Solicited local AEs included injection site pain, redness, swelling, itching. Solicited systemic AEs included fatigue, headache, myalgia, chills, fever, joint pain, nausea, vomiting, diarrhea, abdominal pain, rash external to injection site. We used the Food and Drug Administration AE grading scale ^24^ to classify the severity of self-reported AE.

### Sample collection and processing

Samples were collected during the baseline visit, and at follow-up, 21 days post-dose 1 and -dose 2. Two nasopharyngeal swabs (Med Ams, Russia) were collected following the national guidelines for COVID-19 testing by insertion of the swab into the nasopharynx at a 5-6 cm depth (∼ distance from the nostrils to the outer opening of the ear) for 10 seconds and rotated during slow removal; one swab was then stored for subsequent SARS-CoV-2 PCR testing in DNA/RNA shield media (Zymo Research, Irvine, US), and another in 500 µl of PBS (for nasal Ab measurements). Blood (5 ml) was collected by venipuncture into EDTA tubes (Improvacuter, Gel & EDTA.K2, Improve Medical Instruments, Guangzhou, China). Full blood counts were acquired using a BC3200 automated hematology analyzer (Shenzhen Mindray Bio-Medical Electronics Co., China). Blood plasma was isolated by centrifugation at 2,000×g for 10 minutes. All samples were stored at -80 °C prior to analyses. All readouts were performed using commercially available, well-validated assays deployable in a clinical lab setting ^25–29^. To avoid the bias of inter-run variation, paired (baseline-dose 1-dose 2) samples were examined on the same plate for immunoglobulin, neutralization, and cytokine assays.

### SARS-CoV-2 PCR

Total RNA was isolated from nasopharyngeal swabs by magnetic bead-based nucleic acid extraction (RealBest Sorbitus, Vector-Best, Novosibirsk, Russia) and used for SARS-CoV-2 real-time RT-PCR testing by the Real-Best RNA SARS-CoV-2 kit (Vector-Best, Novosibirsk, Russia) targeting the SARS-CoV-2 RdRp and N loci, following the manufacturer’s protocol.

### Immunoglobulin assays

SARS-CoV-2 S1 IgG, IgA and NCP IgG ELISAs (Euroimmun Medizinische Labordiagnostika AG, Lübeck, Germany) were performed on the Evolis 100 ELISA reader (Bio-Rad) according to the manufacturers’ protocols. Optical density (OD) ratios were calculated as ratio of the OD reading for each sample to the reading of the kit calibrator at 450 nm. For the serologic assays, we used the manufacturer-recommended threshold (0.8) and considered all samples with OD ratios <0.8 and >=0.8 as “negative” and “positive”, respectively. To set the mucosal assay thresholds, we calculated the means and standard deviations of the OD450 ratios of samples from the “no prior COVID-19” subjects at baseline. We then empirically assumed that ∼99.7% of IgG- and IgA-negative samples would fall within 3 standard deviations of the calculated means, i.e. within OD450 ratios of 0.11 and 3.03 for S-IgG and S-IgA, respectively.

### SARS-CoV-2 Surrogate Virus Neutralization Assay

Surrogate neutralization assays (cPass SARS-CoV-2 Neutralization Antibody Detection Kit, #L00847-C, GenScript Biotech Co., Nanjing City, China) were performed by assessing inhibition of the interaction between the recombinant SARS-CoV-2 receptor binding domain (RBD) fragment and the human ACE2 receptor protein (hACE2) ^29^. Briefly, plasma samples and manufacturer-provided controls were pre-incubated with the horseradish peroxidase (HRP)-conjugated RBD at 37 °C for 30 min, and then added to the hACE-2 pre-coated plate for incubation at 37 °C for 15 min. After washing and incubation with the tetramethylbenzidine substrate, absorbance was measured at 450 nm using the Evolis 100 ELISA reader (Bio-Rad). Quality control was performed following the manufacturer’s recommendations. Neutralization % was calculated by subtracting the negative control-normalized absorbance of the samples from 1 and multiplying it by 100%; a manufacturer-recommended cut-off of 30% was used for detectable SARS-CoV-2 neutralization activity.

### Multiplex cytokine ELISA

Blood plasma was analyzed using the Milliplex Map Human Magnetic Bead Panel for cytokines and chemokines (HCYTMAG-60K-PX41) according to the manufacturer’s protocol on a Bio-Plex 3D instrument (Bio-Rad) as done previously ^30^. The ELISA assay details, and analyte classification are given in Appendix Table 1.

**Table 1.** Characteristics of study participants

| Characteristic | Overall, N = 73 | No prior COVID-19, N = 29 | Prior COVID-19, N = 44 | p-value * |
| --- | --- | --- | --- | --- |
| Age, years, median (IQR) | 45.0 (38.0, 58.0) | 44.0 (38.0, 59.0) | 45.5 (38.5, 55.8) | 0.830 |
| Male sex, n (%) | 27 (37.0%) | 9 (31.0%) | 18 (40.9%) | 0.392 |
| Kazakh ethnicity, n (%) | 40 (54.8%) | 11 (37.9%) | 29 (65.9%) | **0.019** |
| BMI, kg/m<sup>2</sup>, median (IQR) | 25.2 (22.8, 27.7) | 23.7 (22.3, 27.5) | 25.5 (24.1, 27.9) | 0.189 |
| Any comorbidities ^ | 35 (47.9%) | 13 (44.8%) | 22 (50.0%) | 0.665 |

\* Differences between the Prior and No Prior COVID-19 groups were assessed using Mann-Whitney U or Pearson's Chi-squared tests.
^ Comorbidities consisted of self-reported gastrointestinal conditions, hypertension, chronic heart disease, chronic obstructive pulmonary disease, history of malignancy, diabetes, liver disease, thyroid dysfunction, kidney disease, neurologic conditions, autoimmune conditions; the distribution of individual comorbidities did not differ between the “no prior COVID-19” and “prior COVID-19” groups.

### Definitions

We defined the “no prior COVID-19” subjects as negative for both S-IgG and -IgA (IgG-, IgA-) and the “prior COVID-19” subjects as positive for either or both IgG and/or IgA (IgG+/-, IgA+/-) at baseline using the serologic S-IgG and S-IgA assay threshold.

### Statistical analysis

We hypothesised that Sputnik-V vaccination in this cohort from Kazakhstan safely boosts both serologic and mucosal SARS-CoV-2-reactive immunity. The primary reactogenicity outcomes were vaccine-elicited self-reported AE. The primary immunogenicity outcomes were vaccine-elicited S-IgG, S-IgA and neutralizing Ab changes. All other clinical and immunologic findings were exploratory outcomes. All analyses were performed in the IBM SPSS V.26 and R V. 4.1.2 (R Core Team, 2021) software. We used the two-sided Mann-Whitney U, Pearson χ2, or Fisher’s exact tests to compare differences between groups, as appropriate and 95% confidence intervals (CI) were calculated using the binomial “exact” method. Reactogenicity between vaccine doses was compared by McNemar’s test. Fold changes were calculated as ratios of geometric mean OD450 ratios or % neutralization, for the serologic Ab-binding and neutralization assays, respectively. Correlations among variables were explored using the Spearman rank test. To facilitate the graphing of geometric means and confidence intervals for the neutralization assay data, “zero” values were substituted with “1”. Cytokine and FBC heatmaps were made by first calculating “follow-up/baseline” ratios of cytokine concentrations for each participant, then by deriving geometric means of individual ratios for each parameter. To minimize the bias contributed by missing data, we focused our analyses on subjects with complete data for at least dose 1 follow-up (N=73).

### Ethics statement

All study procedures were approved by the Research Ethics Board of the Karaganda Medical University. Written informed consent was obtained from all participants.

## RESULTS

### Study participants

We recruited 82 COVID-19-negative adults scheduled to receive their first two Sputnik-V doses (Fig 1). Diagnostic testing was done to identify breakthrough SARS-CoV-2 infection at follow-up visits, since S-specific IgG and IgA tests can not differentiate between immunity acquired by vaccination or natural infection. As a result, there were 4 cases of breakthrough COVID-19 among the participants post-dose 1; no breakthrough infection was detected post-dose 2 (Fig 1). Cumulatively, 73 and 70 participants were sampled during the first and second post-vaccination follow-up visits, respectively (Fig 1, Table 1). The participants were stratified by prior SARS-CoV-2 exposure based on serologically confirmed presence of S-IgG and/or S-IgA [see Methods]. Apart from ethnicity, whereby ethnic Kazakh subjects were over-represented in the “prior COVID-19” group, there were no differences between the subgroups regarding their basic biometric and demographic characteristics (Table 1).

**Fig 1.**
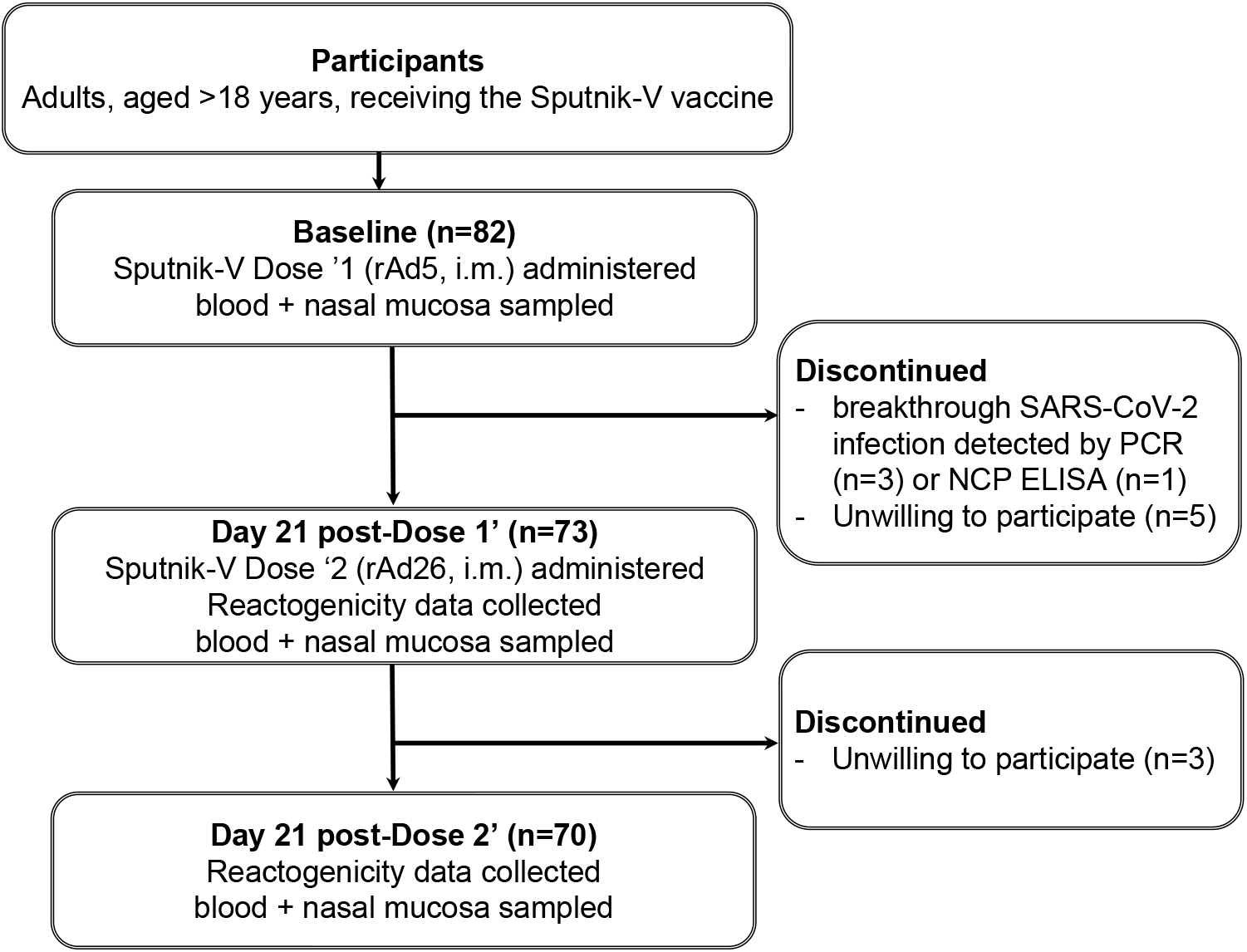
Screening and recruitment flow chart. See the Methods for detailed study inclusion criteria. rAd: recombinant adenovirus; i.m.: intramuscular; NCP: nucleoprotein.

### Sputnik-V-associated adverse events

The self-reported AE associated with Sputnik-V vaccination, a pre-defined primary trial endpoint, were assessed in all participants at 21 days post-dose 1 and at 21 days post-dose 2. All self-reported AE were mild to moderate in nature; we did not detect any severe (grade 3) or potentially life-threatening (grade 4) AE. Most participants reported an injection site (IS) reaction (dose 1: 58%; dose 2: 49%) or a systemic reaction (dose 1: 71%; dose 2: 61%) (Fig 2). The most frequent local and systemic reactions were IS pain (dose 1: 54%; dose 2: 47%), fatigue (dose 1: 53%; dose 2: 47%), myalgia (dose 1: 40%; dose 2: 31%), chills (dose 1: 33%; dose 2: 26%), joint pain (dose 1: 33%; dose 2: 23%), headache (dose 1: 33%; dose 2: 26%) and fever (dose 1: 31%; dose 2: 23%) (Fig 2). Although overall dose 2 appeared to be less reactogenic (Fig 2), there was not a significant difference in reactogenicity between the two doses. In the analysis stratified by prior COVID-19 exposure (Fig 2), fatigue (35% vs. 65%, p=0.016) and headache (44% vs. 17%, p=0.022) were more frequent post-dose 1 in participants with prior COVID-19, while nausea was more common in the “no prior COVID-19” group after dose 2 (14% vs. none, p=0.026). We also noted that participants reporting any IS reactions post-dose ‘1 were more likely to report an IS reaction post-dose ‘2 (Chi square, p<0.001); this was not observed for systemic reactions (p=0.18).

**Fig 2.**
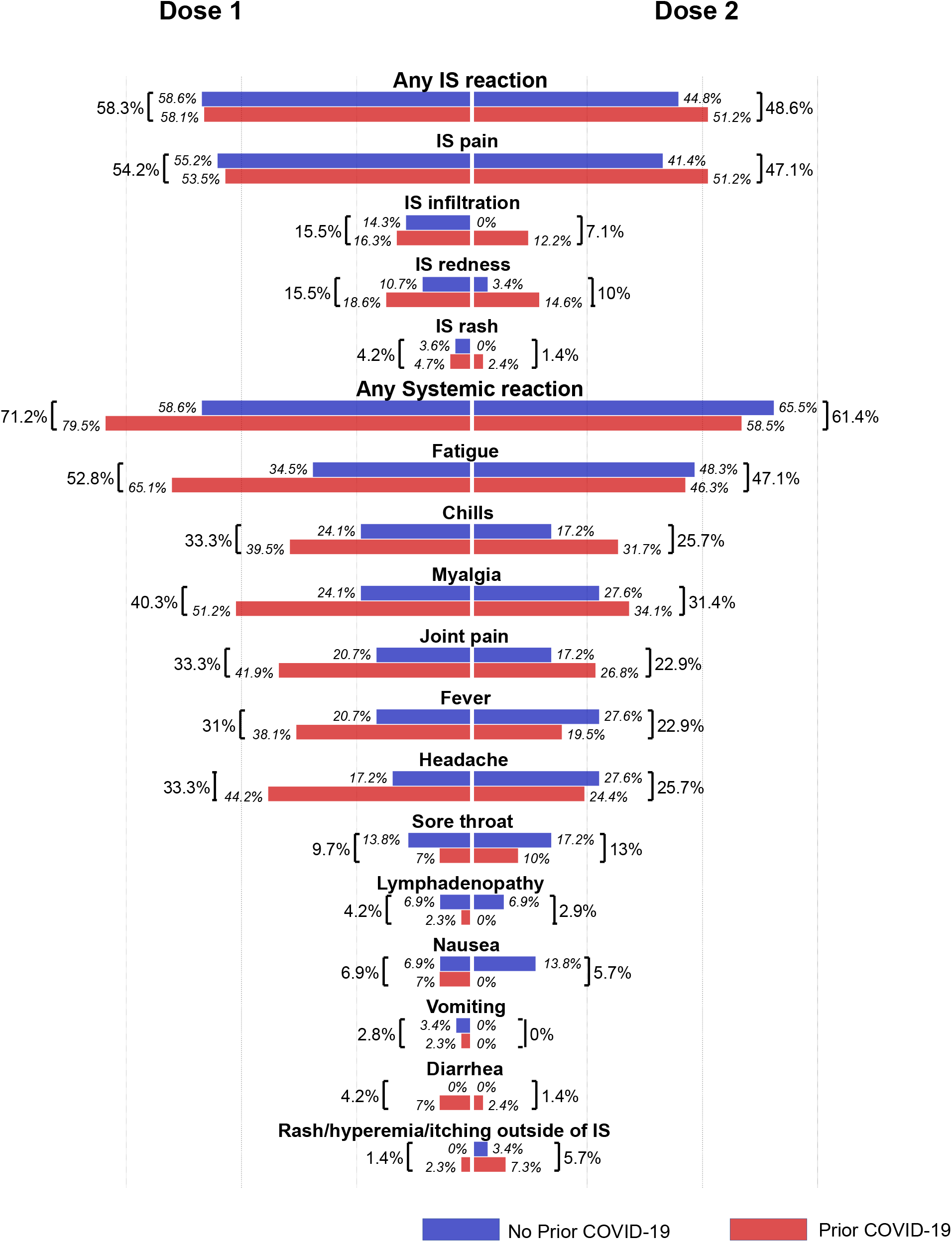
Solicited adverse events (AE) associated with Sputnik -V vaccination. Bars and percentages represent the proportions of participants reporting an AE. IS: injection site.

### Sputnik-V-elicited serologic responses

Titers of Sputnik-V vaccination-induced serologic SARS-CoV-2 S-specific IgG (S-IgG) and IgA (S-IgA), another pre-defined primary trial endpoint, were assessed in all participants at 21 days post-dose 1 and at 21 days post-dose 2 (Figs 3A). S-IgG levels increased 7.7-fold post-dose 1 (p<0.001) and remained high post-dose 2 in all participants (Figs 3A). Compared to dose 1, dose 2 did not further boost S-IgG (p=0.495, Fig 3A). Overall, 90% (66/73) and 91% (64/70) of participants were above the threshold of positivity for S-IgG after dose 1 and dose 2, respectively. Post-dose 1, S-IgA levels increased 5-fold (p<0.001) and remained high post-dose 2 in all participants (Fig 3B); there was no further change after dose 2 compared to dose 1 (Fig 3A). Overall, 93% (68/73) and 89% (62/70) of participants were above the threshold of positivity for S-IgA post-dose 1 and -dose 2, respectively. The combined seroconversion rate based on both S-IgG and S-IgA after dose 1 was 97% (71/73), remaining at 97% (68/70) post-dose 2.

**Fig 3.**
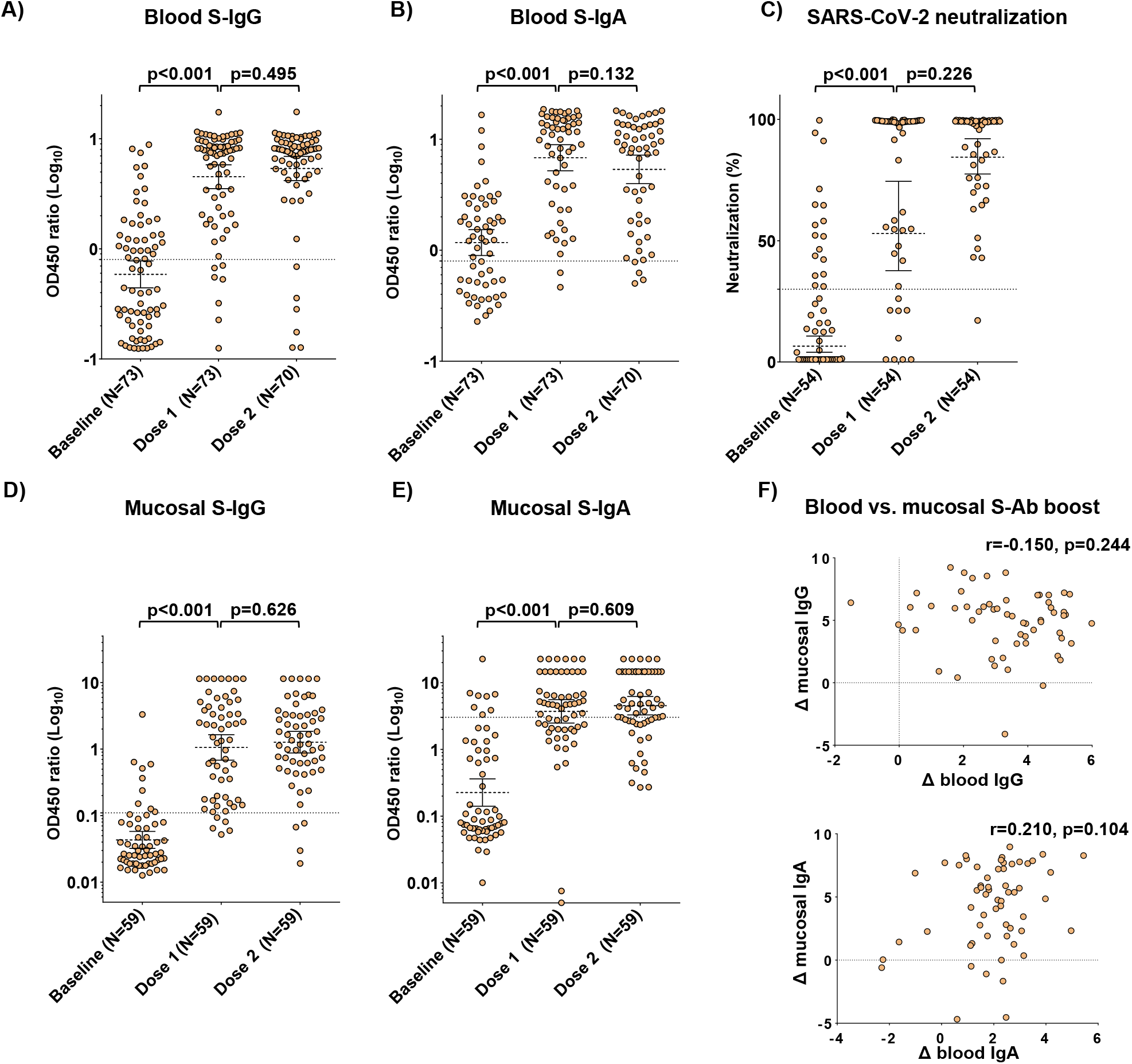
Effects of Sputnik-V vaccination on blood and mucosal SARS-CoV-2 antibodies in all participants. A) Blood Spike-reactive IgG titers at baseline and after vaccination. B) Blood Spike-reactive IgA titers at baseline and after vaccination. C) SARS-CoV-2 neutralization at baseline and after vaccination. D) Mucosal Spike-reactive IgG titers at baseline and after vaccination. E) Mucosal Spike-reactive IgA titers at baseline and after vaccination. F) Correlation plots of mucosal and blood IgG (top) and IgA (bottom) log2-transformed post-dose 2/baseline titer ratios. The Spearman coefficients (r) and their statistical significance (p) are shown. In panels A-E: brackets represent geometric means and 95% confidence intervals; p values indicate the statistical significance assessed by the Mann-Whitney U test. In panels A-E: dotted lines represent the thresholds of assay positivity, defined as OD450 ratio=0.8 for blood S-IgG and S-IgA, SARS-CoV-2 neutralization=30% and OD450 ratio=0.11 and 3.03 for mucosal S-IgG and S-IgA, respectively.

### Sputnik-V-elicited SARS-CoV-2 neutralizing capacity

The effects of Sputnik-V vaccination on the serologic titers of SARS-CoV-2 neutralizing antibodies were next assessed (Fig 3C). Due to limited reagent availability, the assay was performed on a randomly sampled subset of samples (n=54). Virus neutralization increased 8.3-fold post-dose 1 (p<0.001) and remained high post-dose 2 in all participants (Fig3C). Compared to dose 1, dose 2 did not further increase virus neutralization (p=0.226, Fig 3C). Overall, 45/54 (83%) and 53/54 (98%) of tested participants were above the threshold of positivity for neutralization after dose 1 and dose 2, respectively.

### Sputnik-V-elicited mucosal SARS-CoV-2 antibody responses

Mucosal immunity is thought to be important for protection against COVID-19 ^31^. Therefore, we next assessed the S-IgG and S-IgA profiles in nasopharyngeal swab-derived secretions, a secondary trial endpoint, to determine the impact of Sputnik-V on mucosal immunity. Nasal S-IgG levels increased 25-fold after dose 1 (p<0.001) and remained high after dose 2 in all participants (Fig3D). Compared to dose 1, dose 2 did not further increase IgG levels (p=0.626, Fig3D). Nasal IgA levels increased 16.5-fold after dose 1 (p<0.001) and remained high after dose 2 in all participants (Fig3E). Compared to dose 1, dose 2 did not further increase IgA levels (p=0.609, Fig3E). The magnitudes of S-IgG and S-IgA responses did not correlate between the blood and mucosa (Fig 3F and 4F).

**Fig 4.**
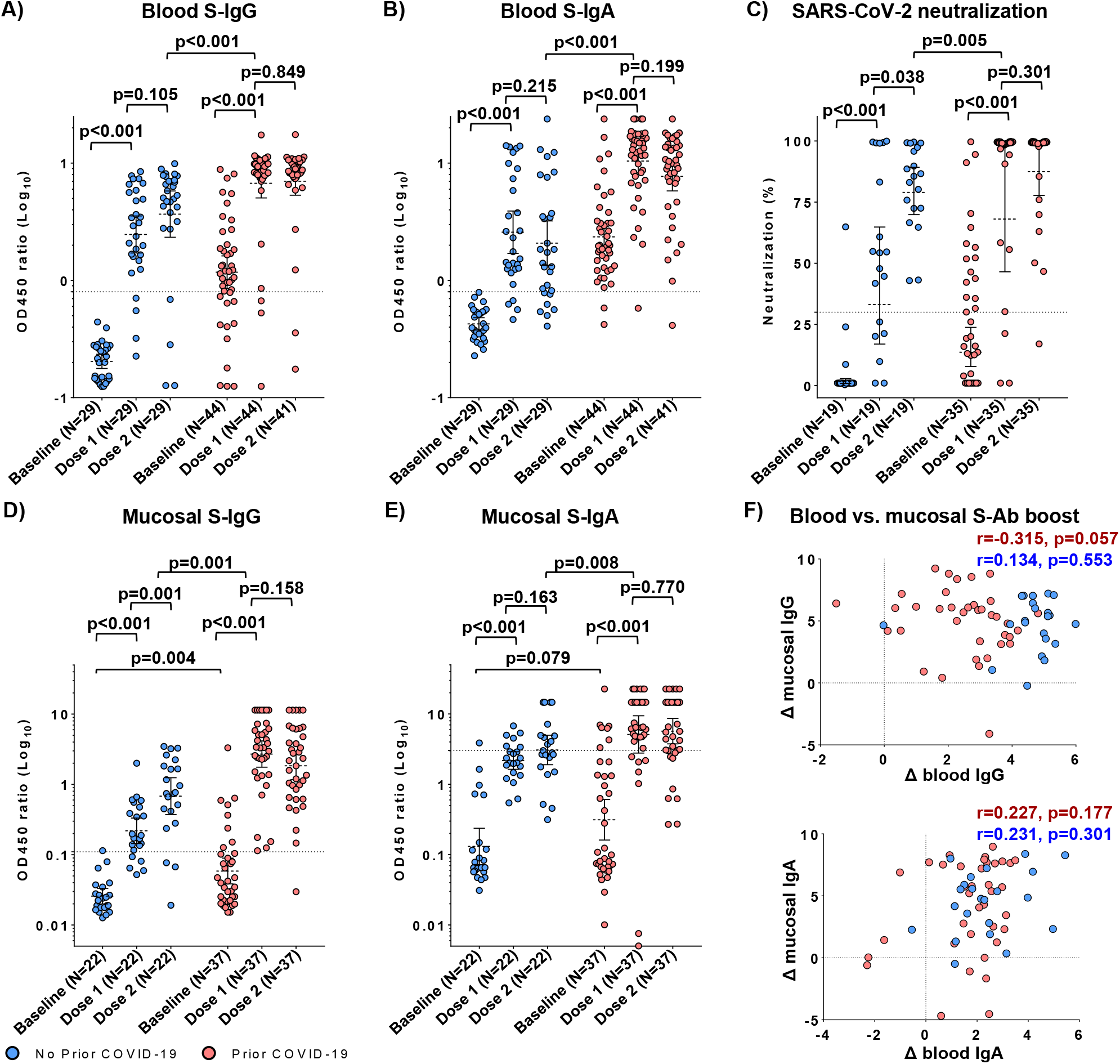
Effects of Sputnik-V vaccination on blood and mucosal SARS-CoV-2 antibodies in participants stratified by prior exposure to COVID-19. A) Blood Spike-reactive IgG titers at baseline and after vaccination. B) Blood Spike-reactive IgA titers at baseline and after vaccination. C) SARS-CoV-2 neutralization at baseline and after vaccination. D) Mucosal Spike-reactive IgG titers at baseline and after vaccination. E) Mucosal Spike-reactive IgA titers at baseline and after vaccination. F) Correlation plots of mucosal and blood IgG (top) and IgA (bottom) log2-transformed post-dose 2/baseline titer ratios. The Spearman coefficients (r) and their statistical significance (p) are shown. In panels A-E: brackets represent geometric means and 95% confidence intervals; p values indicate the statistical significance assessed by the Mann-Whitney U test. In panels A-E: dotted lines represent the thresholds of assay positivity, defined as OD450 ratio=0.8 for blood S-IgG and S-IgA, SARS-CoV-2 neutralization=30% and OD450 ratio=0.11 and 3.03 for mucosal S-IgG and S-IgA, respectively.

### Effects of prior COVID-19 exposure on Sputnik-V-elicited immunity in blood

Earlier studies have that individuals with prior COVID-19 exposure generated a stronger immune response after a single vaccine dose than naïve individuals ^16–18,32^. To test whether this held true in our cohort, we compared Sputnik-V-elicited SARS-CoV-2 immune responses in the ‘prior COVID-19” and “no prior COVID-19” groups in the blood (Fig 4A-C) and mucosa (Fig 4D-E).

While dose 1 increased the titres of serologic S-IgG and S-IgA, dose 2 did not have a perceivable boosting effect on either S-IgG or S-IgA (Fig4 A-B). Compared to the “prior COVID-19” subjects post-dose 1, S-binding Ab titres were lower in the “no prior COVID-19” group post-dose 2 (1.8- and 5-fold, p<0.001 for IgG and IgA, respectively, Fig 4A-B).

Whereas neutralization was robustly boosted by dose 1 in both the “prior” and “no prior COVID-19” groups, dose 2 was associated with an increase in neutralization only in the “no prior COVID-19” group (2.4-fold; p=0.038) (Fig4C). Notably, compared to the “prior COVID-19” subjects post-dose 1, neutralizing antibody titers were 1.2-fold higher in the “no prior COVID-19” group (p=0.005) post-dose 2 (Fig. 4C). Thus, overall in this cohort prior COVID-19 exposure was associated with stronger vaccine-elicited serologic responses, with effects on the binding abs slightly more pronounced compared to neutralizing Ab titres.

### Effects of prior COVID-19 exposure on Sputnik-V-elicited immunity in mucosa

Subjects with prior COVID-19 exposure had higher mucosal S-IgG compared to the “no prior COVID-19” group (p=0.004) (Fig 4D). Vaccination-elicited boost of mucosal IgG in the “prior COVID” participants was higher in magnitude (45.8 and 31.2-fold for dose 1 and 2’, respectively) compared to the “no prior COVID-19” subjects (8.4 and 26.2-fold for dose 1 and 2’, respectively, Fig 4D).

Baseline mucosal S-IgA titres also tended to be higher in the “prior COVID-19” participants, although this difference was not significant (p=0.079, Fig 4E). Vaccination-elicited mucosal S-IgA boost after each dose was similar in magnitude in participants with prior COVID-19 (16.6 and 23.5-fold for dose 1 and 2’, respectively) compared to those without prior COVID-19 (16.4 and 18.2-fold for dose 1 and 2’, respectively, Fig 4E). Consistent with the unstratified analysis, the magnitudes of S-IgG and S-IgA responses did not correlate between the blood and mucosa in the “prior” vs. “no prior” COVID groups (Fig 4F).

Compared to the “no prior COVID-19” subjects post-dose 2, both mucosal S-IgG and S-IgA titres were higher (4.0 and 1.7-fold, p=0.001 and p=0.008, respectively) in the “prior COVID-19” group after a single Sputnik-V dose (Fig 4D-E). Overall, in this cohort prior COVID-19 exposure was associated with a stronger vaccine-elicited mucosal immunity.

### Effects of Sputnik-V on systemic immunology

Lastly, we explored the impact of Sputnik-V vaccination on cytokine changes in the blood using a multiplex assay targeting 41 analytes (see Appendix for complete list). We observed that the only analyte significantly affected by vaccination was growth regulated oncogene (GRO), the concentration of which post-dose 1 and 2’ was reduced by 2.0-(p = 0.007) and 2.2-fold (p = 0.003), respectively (Fig 5A-B), while notable trends were also seen for some chemokines (eotaxin, RANTES and IP-10) and proinflammatory cytokines TNF-a and IL-17 in association with dose 2 (Fig 5A). Changes in GRO concentration did not correlate with S-IgG or S-IgA changes (Appendix Fig S1). We therefore hypothesized that Sputnik-V-elicited reduction in GRO concentration might be related to alterations in some circulating cells type(s). To test this hypothesis, we first analyzed full blood counts (FBC), which were available at both baseline and post-vaccination follow-up for a subset of participants (N=34, see Appendix Table S2), and then assessed whether the GRO change was associated with FBC alterations. Counts of all assessed cell types, including granulocytes, lymphocytes, monocytes, platelets, and erythrocytes, were elevated 21 days after both vaccine doses (Fig 5C). Absolute platelet counts were inversely correlated with Sputnik-V-induced GRO reduction (Fig 5D). Notably, at baseline 24% (8/34) of the subjects had mild thrombocytopenia (median platelet count=122×10^9^ cells/L), while at follow-up no thrombocytopenia was detected and one subject had thrombocythemia (511×10^9^ cells/L), highlighting a clinically meaningful increase to circulating platelet counts elicited by Sputnik-V.

**Fig 5.**
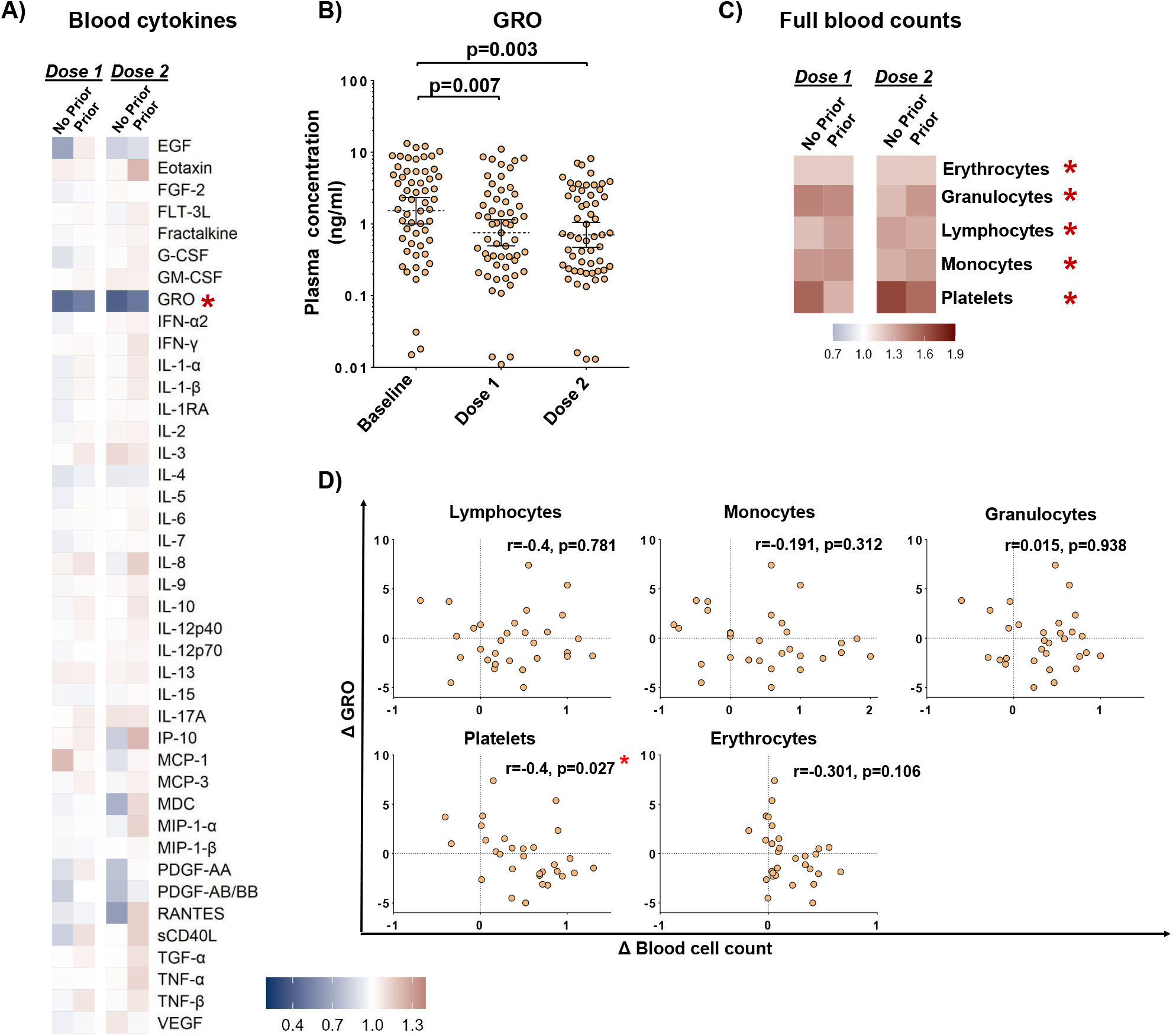
Systemic cytokine and cellular changes associated with Sputnik-V vaccination. A) Blood cytokine changes represented as the geometric mean of fold change of each cytokine post-vaccination (after dose 1 and 2) over pre-vaccination level in the No Prior (N=21) and Prior (N=36) COVID-19 groups. Cytokines were plotted in alphabetical order (see the Methods and Appendix for the details). B) Blood growth regulated oncogene (GRO) concentration at baseline and after Sputnik-V vaccination in the No Prior (N=21) and Prior (N=36) COVID-19 groups. Brackets represent geometric means and 95% confidence intervals; p values indicate the statistical significance assessed by the Mann-Whitney U test. C) Changes in full blood counts (FBC) represented as the geometric mean of fold change of each cell sub-type post-vaccination (after dose 1 and 2) over pre-vaccination count in the No Prior (N=10) and Prior (N=25) COVID-19 groups. D) Correlation plots of blood GRO (y-axis) and FBC-derived counts of major blood cell subtypes (x-axis). Post-dose 2/baseline ratios were log2-transformed. The Spearman coefficients (r) and their statistical significance (p) are shown. In panels A, C: Scale denotes fold changes, whereby 1.0= “no change”. Stars represent statistically significant differences before and after vaccination at p<0.05.

## DISCUSSION

Here we assessed the reactogenicity and immunogenicity of Sputnik-V in a prospective clinical trial involving subjects with and without prior COVID-19 exposure in Kazakhstan. The observed reactogenicity was similar to that reported by earlier studies of Sputnik-V ^1,2,5–9^ and other COVID-19 vaccines ^33^. Consistently, systemic side-effects were more frequent in subjects with prior COVID-19 history. The high SARS-CoV-2 seroconversion rates observed post-vaccination also align well with studies of both Sputnik-V and other COVID-19 vaccines ^1,2,4,7,8,10–12,15–18^. We further show that both serologic and nasal S-IgG and S-IgA are effectively boosted by the Sputnik-V prime shot. Importantly, the effects of dose 2 were limited to neutralizing abs and mucosal S-IgG only in participants without prior COVID-19 history, consistent with prior studies ^16,18,34^. The Sputnik-V-elicited alterations to the GRO/platelet axis call for further investigation of Sputnik V effects on systemic immunology.

The current deployment of Sputnik-V is occurring despite the scarcity of research on the safety and immunologic characteristics across different populations. Although this was justifiable at the early stages of the pandemic, more detailed data are urgently needed to inform the immunization programs across different nations in the wake of the rapidly evolving COVID-19 dynamics.

Notably, the unique, heterologous design of Sputnik-V was proposed by the vaccine creators to avoid immunogenicity issues stemming from anti-vector immunity ^1,2^. Our findings support the earlier reports that Sputnik-V has a similar safety and immunogenicity profile to other approved AdV- and mRNA-based vaccines. Furthermore, our finding that prior COVID-19 history is linked to a higher likelihood of vaccination-elicited systemic adverse events is also in keeping with other COVID-19 vaccine studies ^33^.

The high seroconversion rate, calculated based on both S-IgG and S-IgA in our cohort, is similar to earlier studies of Sputnik-V ^2,7,16–18^, and consistently, there was little benefit of Sputnik-V dose 2 - limited to neutralizing Ab and mucosal S-IgG in participants without prior COVID-19. We also observed that Sputnik-V dose 1 boosted both S-IgA and S-IgG in blood and mucosa. This is remarkable given the emerging evidence that serologic S/RBD-specific IgA was associated with protection against breakthrough COVID-19 in mRNA vaccinees ^19^. Consistent with the latter, not only did Sputnik-V dose 2 not boost systemic or mucosal S-IgA, but S-IgA also tended to decline post-dose 2.

Our data suggest that adjustments to the current vaccination regimen interval may be necessary to optimize the immune effects of Sputnik-V dose 2. In support of this, recent studies of mRNA and AstraZeneca vaccines have demonstrated that extending the interval between the doses results in more efficient boosting of antibody titres ^35–37^. Hence, a similar increase of the inter-dose interval would be expected to result in a stronger immunity elicited by the Sputnik-V second dose.

We observed that in the mucosa Sputnik-V elicited a substantial Ab boost, with a more prominent effect on S-IgG compared to S-IgA. This finding corroborates the recent data that intramuscularly administered COVID-19 vaccines can impact mucosal immunity ^19,34,38,39^. Thus, comparisons of mRNA and inactivated virus (CoronaVac) vaccines, indicated that mRNA-based Comirnaty (Pfizer-BioNTech) but not CoronaVac induced S-IgG and -IgA in the nasal epithelial lining fluid ^38^, while mRNA vaccines also boosted S-abs in the saliva ^19,34,39^. Furthermore, mRNA vaccinees with prior COVID-19 exposure mounted stronger salivary S-IgA responses compared to COVID-19-naive vaccinees ^34^. Although the implications of this for protection against COVID-19 are unclear, nasal IgA contributes to protection against human influenza ^40,41^ and SARS-CoV-2 in mice ^42^. Similar to our findings in influenza-vaccinated subjects ^43^, in this cohort we did not observe a correlation between Ab responses in the blood and nasal mucosa, suggesting that vaccine-elicited systemic immunity may not be assumed to accompany mucosal immunity, and emphasizing a potential benefit of mucosal sampling in clinical trials.

Our systemic cytokine analysis indicated subtle effects of Sputnik-V on most of the assessed soluble mediators in blood at 21 days post-vaccination, consistent with the data indicating that mRNA-based Comirnaty-altered cytokine responses also return to the baseline pre-vaccination levels by day 22 post-vaccination ^44^. In the light of this, the sustained reduction of systemic GRO sustained for at least 21 days after Sputnik-V vaccination seen in our study was somewhat unexpected. The GRO family of chemokines consists of GRO-α, GRO-β and GRO-γ or chemokine (C-X-C motif) ligands (CXCL) 1, 2 and 3, respectively, with GRO-α representing the bulk of circulating GRO (∼80%) ^45^. All three chemokines share one receptor, CXCR2 (C-X-C Motif Chemokine Receptor 2), and have been implicated in responses to viral infection, hematopoietic malignancies and vascular disease ^46–49^. Innate immune cells, including monocytes^49^ and neutrophils ^45^, and platelets ^50^ have been identified as major GRO producers. Post-vaccination elevations of major blood cell subtypes is not surprising, however, it is intriguing that vaccine-induced GRO reduction was correlated with elevated platelet counts. Alterations in platelet functionality have been proposed to play a role in the rare immune thrombocytopenia events associated with COVID-19 vaccines (but has not been reported for Sputnik-V) ^4^. Therefore, the observed alterations to the GRO/platelet axis in the context of viral vectored vaccines could provide insight into the mechanism of vaccine-driven alterations of platelet dynamics.

Important limitations of our study are the relatively small sample size and short duration of post-vaccination follow-up, although these limitations were partially overcome by the longitudinal design with a high retention (>85%) of participants throughout the study follow-up. Our cohort was also female-dominated, which may have biased the results of both SAE and immunologic outcomes.

In summary, our study contributes to the growing body of evidence that Sputnik-V vaccination safely elicits robust immune responses, both serologically and mucosally. Our findings also suggest that the current Sputnik-V dosing interval guidelines need urgent adjustment given the minimal benefit observed after dose 2. Finally, the impact of Sputnik-V on platelets merits further investigation to better our understanding of rare adverse events related to thrombosis and thrombocytopenia that have been reported for COVID-19 vaccines.

## Data Availability

All data produced in the present study are available upon reasonable request to the authors

## CONTRIBUTORS

Conceptualization, SY, IK, DB. Data curation, SY, IlK, DB. Investigation and formal analysis, SY, IK, BN, YK, SK, IlK, YB, BM, MSM, MSM, GH, DB. Clinical and laboratory site supervision IK, BN, BM, GH. Writing – original draft, SY, IK, MSM, DB. Writing – review and editing, SY, IK, BN, YK, SK, IlK, YB, BM, MSM, MSM, GH, DB. Funding acquisition, IK, BM, GH, DB.

## ACKNOWLEDGEMENTS

We thank all the participants and clinical and laboratory staff, who have been involved in the study.

## DATA SHARING

The source data and R code used for all analyses will be made available through the Github.com server upon article acceptance. Any additional data from this study will be made available, wherever possible, on appropriate request to the corresponding author.

## SUPPLEMENTARY INFORMATION

All supplementary information can be found in the Appendix.

