## Appendix for "Sputnik-V reactogenicity and immunogenicity in the blood and mucosa: a prospective cohort study"

*This supplemental material has been provided by the authors to give readers additional information about their work.*

**Supplementary Table 1.** Description of the analytes assessed by multiplex ELISA.

| **#** | **Analyte** | **Functional category** | **Full name** | **Alternative name** | **Lowest limit of detection (pg/ml)** |
| --- | --- | --- | --- | --- | --- |
| 1 | IL-1α | Proinflammatory | Interleukin-1α | - | 9.4 |
| 2 | IL-1β |  | Interleukin 1β | - | 0.8 |
| 3 | IL-6 |  | Interleukin 6 | - | 0.9 |
| 4 | TNF α |  | Tumor necrosis factor α | DIF | 0.7 |
| 5 | TNF β |  | Tumor necrosis factor-β | Lymphotoxin-α (LT-α) | 1.5 |
| 6 | IL-17A |  | Interleukin 17a | CTLA8 | 0.7 |
| 7 | sCD40L |  | CD40 ligand | IGM; IMD3; TRAP; CD154; | 5.1 |
| 8 | IL-2 | Homeostatic | Interleukin 2 | TCGF | 1.0 |
| 9 | IL-7 |  | Interleukin 7 | - | 1.4 |
| 10 | IL-12p40 | Th1/Th2 | Interleukin-12 p40 | Interleukin-23 | 7.4 |
| 11 | IL-12P70 |  | Interleukin 12 p70 | CLMF; NKSF; CLMF2; IMD28; IMD29; NKSF2; | 0.6 |
| 12 | IL-4 |  | Interleukin 4 | BSF1 | 4.5 |
| 13 | IL-5 |  | Interleukin 5 | EDF; TRF | 0.5 |
| 14 | IL-13 |  | Interleukin 13 | P600 | 1.3 |
| 15 | IL-9 | Th9 | Interleukin 9 | P40; HP40 | 1.2 |
| 16 | IL-1Ra |  | Interleukin-1 receptor antagonist | DIRA; IRAP; IL1F3; IL1RA; MVCD4; | 8.3 |
| 17 | IL-10 | Anti-inflammatory | Interleukin 10 | CSIF; TGIF; GVHDS | 1.1 |
| 18 | TGF-α |  | Transforming growth factor α | - | 0.8 |
| 19 | IFN-α2 | Interferons | Interferon α2 | - | 2,9 |
| 20 | IFN-γ |  | Interferon γ | IFG; IFI | 0.8 |
| 21 | IL-3 | Growth factors | Interleukin 3 | MCGF; MULTI-CSF | 0.7 |
| 22 | IL-15 |  | Interleukin 15 | - | 1.2 |
| 23 | EGF |  | Epidermal growth factor | URG; HOMG4 | 2,8 |
| 24 | FGF-2 |  | Fibroblast growth factor 2 | BFGF; FGFB; HBGF-2 | 7,6 |
| 25 | PDGF AA |  | Platelet-derived growth factor AA | PDGF1 | 0.4 |
| 26 | PDGF AB/BB |  | Platelet-derived growth factor AB/BB | PDGF2 | 2.2 |
| 27 | VEGF A |  | Vascular endothelial growth factor A | VPF; MVCD1 | 26.3 |
| 28 | Fit-3L |  | Fms-related tyrosine kinase 3 ligand | CD135 | 5.4 |
| 29 | G-CSF | Colony-stimulating factors | Granulocyte-colony stimulating factor | Colony-stimulating factor 3 | 1.8 |
| 30 | GM-CSF |  | Granulocyte-macrophage colony-stimulating factor | Colony-stimulating factor 2 | 7.5 |
| 31 | IL-8 | Chemokines | Interleukin 8 | CXCL8 | 0.4 |
| 32 | Eotaxin |  | Eosinophil chemotactic protein | CCL11 | 4,0 |
| 33 | Fractalkine |  | Fractalkine | CX3CL1, neurotactin | 22.7 |
| 34 | GRO |  | human growth-regulated oncogene | CXCL1-3 | 9.9 |
| 35 | IP-10 |  | Interferon gamma-induced protein 10 | CXCL10 | 8.6 |
| 36 | MCP-3 |  | Monocyte-chemotactic protein 3 | CCL7 | 3.8 |
| 37 | MCP-1 |  | Monocyte-chemotactic protein 1 | CCL2 | 1.9 |
| 38 | MDC |  | Macrophage-derived chemokine | CCL22 | 3.6 |
| 39 | MIP-1α |  | Macrophage inflammatory protein 1-α | CCL3 | 2.9 |
| 40 | MIP-1β |  | Macrophage inflammatory protein 1- β | CCL4 | 3.0 |
| 41 | RANTES |  | Regulated on activation, normal t cell expressed and secreted | CCL5 | 1.2 |

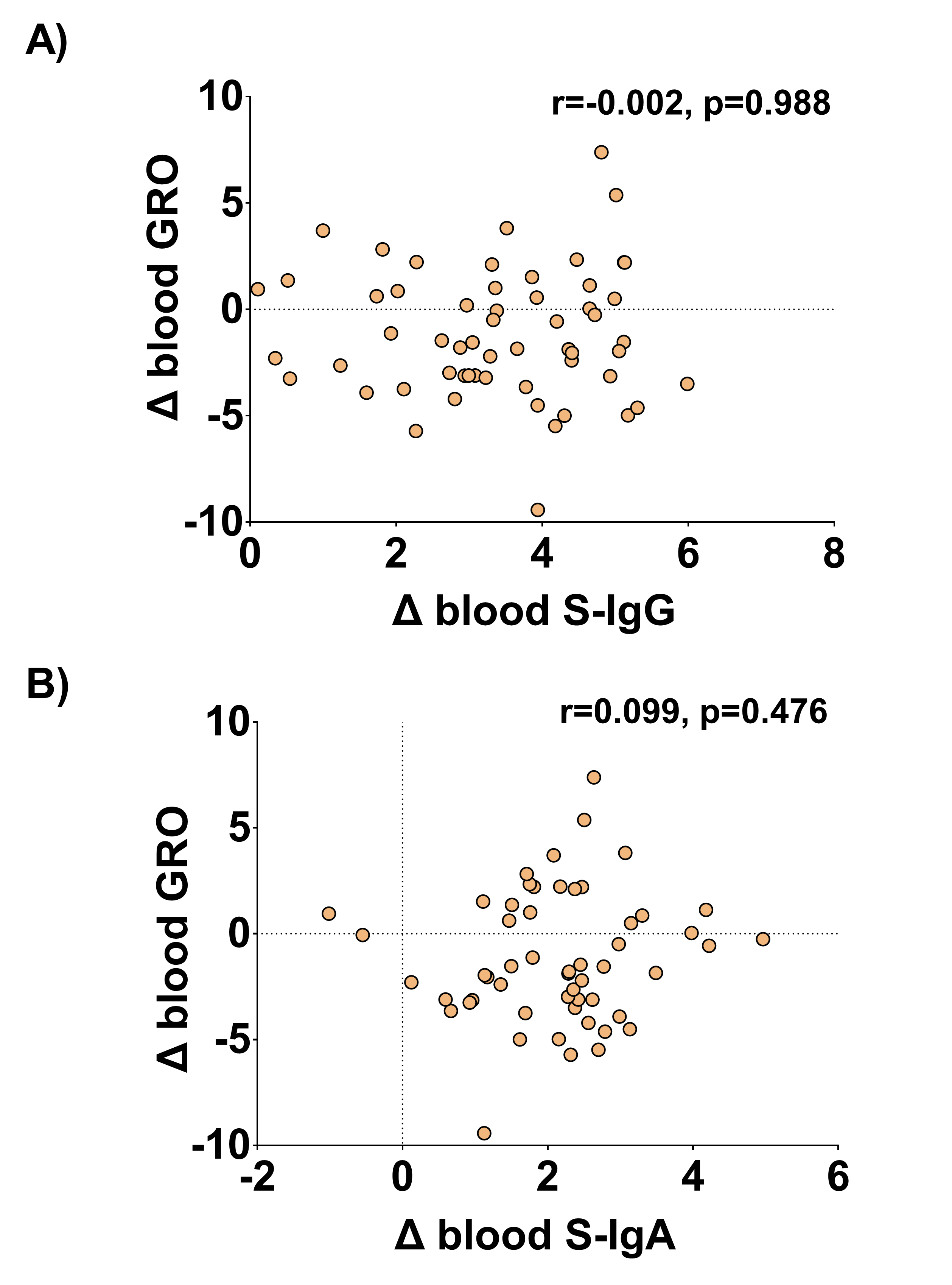

**Figure S1.** Correlation plots of blood GRO (y-axis) and blood S-IgG (top) and S-IgA (bottom) (x-axis) ratios. Post-Dose 2/baseline ratios were log2-transformed. The Spearman coefficients (r) and their statistical significance (p) are shown.

**Table S2.** Characteristics of participants with full blood count data available for both baseline and post-dose 2’ visits.

| **Characteristic** | **Overall, N = 34** | **No prior COVID, N = 9** | **Prior COVID, N = 25** | **p-value^*^** |
| --- | --- | --- | --- | --- |
| Age, years, median (IQR) | 44.0 (37.8, 53.0) | 40.0 (37.0, 52.0) | 44.0 (41.0, 53.0) | 0.598 |
| Male sex, n (%) | 12 (35.3%) | 2 (22.2%) | 10 (40.0%) | 0.439 |
| BMI, kg/m^2^, median (IQR) | 24.8 (22.9, 25.9) | 23.7 (22.5, 28.8) | 25.1 (24.0, 25.8) | 0.648 |
| Kazakh ethnicity, n (%) | 20 (58.8%) | 4 (44.4%) | 16 (64.0%) | 0.435 |
| Any comorbidities | 16 (47.1%) | 4 (44.4%) | 12 (48.0%) | >0.999 |

* Differences between the Prior and No Prior COVID-19 groups were assessed using Mann-Whitney U or Pearson's Chi-squared tests.

**Table S3.** List of reactogenicity and immunogenicity studies of Sputnik-V as of 13 January 2022.

| **Outcomes** | **Countries** | **References** |
| --- | --- | --- |
| **Post-vaccination adverse events** | Argentina, Bosnia and Herzegovina, Iran, Russia, Serbia, San-Marino, Venezuela | ^1–7^ |
| **Vaccine-elicited immunologic changes:** |  |  |
| S/RBD-specific ab titres | Argentina, Bosnia and Herzegovina, Iran, Pakistan, Russia, Serbia, Sri Lanka, Venezuela | ^1,2,5,6,8–16^ |
| Other: systemic cytokines | Russia | ^11^ |
| Other: cellular assays/immunospot | Russia, Sri Lanka | ^1,2,10^ |
